# Impact of Musical Therapeutic Intervention on Depression in Dementia Patients and Caregiver Burden

**DOI:** 10.1101/2025.07.17.25325681

**Authors:** Hieu D. Nguyen, Tomoteru Seki, Wendy Chase

**Author notes:** Corresponding Author: Hieu D. Nguyen, A.A, Department of Psychiatry and Behavioral Sciences, Stanford University, Stanford, CA.

## Abstract

Among people with Dementia, those with depression have worse outcomes in quality of life and mortality. More research is needed on non-pharmacological interventions on those with Dementia and their caregivers. Methods: This study investigates receptive music therapy as a non-pharmacological treatment, which has demonstrated previous improvements in depression among those with Dementia and their caregivers. This study utilizes a paired sample t-test to compare patients’ and caregivers’ depression and burden, respectively, before the intervention and after. Results: Findings indicate that music intervention substantially improved Geriatric Depression scores after the music intervention for patients (Cohen’s D = 1.88) and Caregiver Burden scores (Cohen’s D = 0.82). Conclusions: These results demonstrate the impacts of receptive music therapy on depression among Dementia patients and caregivers and demonstrates the need for its continual research to incorporate this treatment into the clinical setting.

## Background

Dementia is a term that encompasses diseases that progressively affect a person’s cognitive function, such as memory.^1^ There are currently over 55 million people worldwide who have dementia.^2^ Dementia is the 7^th^ leading cause of death among all diseases, and Alzheimer’s is the most common form of dementia accounting for 60-70% of all cases.^2^ During this cognitive decline, these individuals will experience a broad range of symptoms, or behavioral and psychological symptoms of dementia (BPSD), which can include depression. Depression is characterized by symptoms such as anhedonia, fatigue, low mood, and changes in eating and sleeping patterns.^3–4^ The prevalence of depression is reported to be highest in those who have mild cognitive impairment.^5^ Those with both cognitive impairment and depression have associated increased rates of mortality and decreased quality of life.^6–7^ This can also increase caregiver burden.^8^ Pharmacological and non-pharmacological interventions have been utilized to treat dementia symptoms. Common pharmacological treatments for BPSD are sedatives and antidepressants. Early and long-term use of SSRIs may delay the risk of developing dementia.^9^ However, antidepressants may not be currently effective in treating depression in patients with dementia.^10^ More studies are needed to form a consensus. In the clinical setting, pharmacological treatments are used much more than non-pharmacological interventions although non-pharmacological interventions are recommended to be the first treatment for BPSD.^11–12^ Although a pharmacological regimen remains the current treatment of choice, it does not provide a comprehensive intervention to fully treat the progression and symptoms of dementia. Though there is much less research on non-pharmacological treatments, recent findings indicate that adding music to standard treatment protocol is associated with benefits in mood for both dementia patients as well as caregivers.^1^ Music therapy has been used in many ways to treat those with depression. In receptive techniques, music is precomposed and played for patients, which can bring relaxation and changes in mood.^12^ Caregivers may also experience caregiver burden while providing significant physical and emotional duties to patients. Caregiver burden has been associated with reduced physical health, and quality of life and can be predictive of anxiety and depression.^13–17^ These outcomes may also result in increased morbidity and mortality.^18–19^ It is important to support caregivers at risk for increased levels of burden to improve their quality-of-life outcomes and quality of care. While Music therapy can alleviate depression and other end-of-life symptoms, very little research has been done to investigate and demonstrate the feasibility of music therapy on caregivers.^20^ In this study, we investigate whether live music interventions have any impact on dementia patients’ depression and caregivers’ burden.

## Methods

### Participants

The study took place at a 32-acre retirement community (Amavida) in Southwest Florida that houses 460 residents of independent living, assisted living, and memory care. Participants were recruited from the Memory Care facility, which offers specialized care to patients with Alzheimer’s disease or other forms of dementia according to the criteria of the Diagnostic and Statistical Manual IV.^21^ Caregivers and physicians at the residence provided optimal personal care to patients with behavioral and psychologic symptoms (BPSD) of dementia.^22^

### Inclusion and Exclusion Criteria

Inclusion Criteria: included residents were over 60 years of age at the facility in the Memory Care Unit, who had been diagnosed with Alzheimer’s disease and had completed cognitive function tests at the facility. Any caregiver at the facility over 25 years of age with at least a year of experience was included in the study.

Exclusion criteria: included patients who were severely impaired (<17) or could not give coherent answers to the MMSE.

### Intervention

Using a receptive music therapy approach, six 30-minute live piano performances were implemented consistently one hour before dinner in front of patients and caregivers over the course of 2 weeks.^23^ The same songs are played in the same order in each session. During the recording, caregivers were instructed to stand in the back, to not cause any disturbance unless patients showed signs of distress.

### Data Collection

Upon Florida SouthWestern State College Institutional Review Board (IRB) approval, in-house Mini-Mental Status Exam (MMSE) data was collected to establish residents’ baseline cognitive functions.^24^

Geriatric Depression Scale (GDS) and Caregiver Burden Scale in End-of-Life Care (CBS-EOLC) were administered before and after music intervention sessions. Caregivers and nurses administered the GDS to patients and evaluated their answers. Participant MMSE scores ranged from 17-30. Any patient in the memory care unit were welcomed to listen to the performance. Following the piano session, any patients who volunteered to be a part of the study would complete a consent form with the help of their caregiver.

Scores are collected from the caregivers and then entered into an Excel sheet at the facility. The patients were assured of their confidentiality in the study and that they could withdraw from the study or refuse answering questions at any time. Data collection began after the participants provided signed informed consent.

### Questionnaires

The MMSE was used to assess patients’ cognitive impairment. The MMSE has been widely accepted among health professionals who oversee those with dementia. It reports a total score of 30 and is categorized into severe cognitive impairment, moderate cognitive impairment, mild cognitive impairment, and may be normal with no cognitive impairment. Its use for screening mildly cognitively impaired patients has been well demonstrated.^25^ Its validity in screening for dementia and mild cognitive impairment is established and also makes it a standard of comparison for other tools.^26^

The Short Form GDS used consisted of 15 questions, which indicates mild, moderate, and severe depression. It has been used as an effective screening tool to manage depression in older adults, and its validity and reliability have been demonstrated in the literature and clinical settings.^27^

Using the CBS-EOLC, we measured caregivers’ perceived emotional and physical exhaustion, demands of care, and locus of control in the clinical setting.^28–29^This self-report, 16-item questionnaire uses a 4-point Likert Scale that scores from a total of 16 to 64. The scale has proven reliability and convergent validity with depression and exhaustion.^28^

### Statistical Analysis

12 patients’ pre- and post-GDS scores were recorded and analyzed. Caregivers pre- and post-Burden Scores were also analyzed. All statistical analysis was conducted through R Studio 4.2.2 for Windows.

A Shapiro-Wilk normality test is used to assess the normality of the data. The data follows a normal distribution, so a paired t-test is used to compare the GDS scores. P-values < 0.05 are considered statistically significant.

## Results

Of the 40 residents who attended the Music Therapy sessions, 12 completed the pre- and post-GDS. Demographic data were collected for both residents and caregivers. The age of residents ranged from 68 to 74 years for males (n=5) and 67 to 82 for females (n=7), with an average age of 70 for males and 74 for females (Table 1). Among the caregiver sample (n=13), age ranged from 29 to 31 years for males (n=3) and 35 to 50 for females (n=10), with an average age of 30 for males and 42 for females (Table 1).

**Table 1.** Age Range and Average Age of Male and Female Residents and Caregivers.

| Residents | Total (n=12) | Age Range | Average Age |
| --- | --- | --- | --- |
| Male | 5 | 68-74 | 70 |
| Female | 7 | 67-82 | 74 |
| Caregivers | Total (n=13) |  |  |
| Male | 3 | 29-31 | 30 |
| Female | 10 | 35-50 | 42 |

Of the residents who did not participate, many were severely cognitively impaired, had impaired vision, and 2 refused. Only 13 residents filled out the initial questionnaire, and 12 of them finished the study, with only 1 dropping out due to personal conflicts. All caregivers completed the initial questionnaire but three dropped out due to unknown reasons. In total, 13 of 16 caregivers completed the initial and post-intervention CBS-EOLC. The average baseline MMSE mean score is 22, indicating mild cognitive impairment among the residents (Table 2).

**Table 2.**
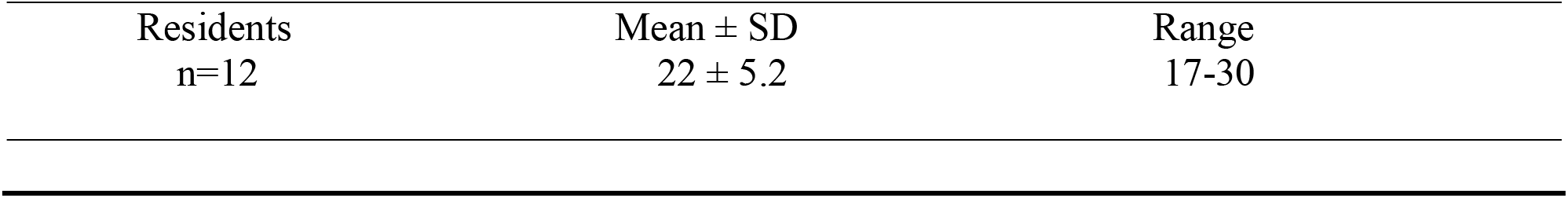
MMSE Scores for Residents at Baseline.

The average GDS score for residents before receiving the music intervention was 3.75 (no depression) with a standard deviation of 1.75. The mean GDS score for residents after receiving the music intervention was 0.75, with a standard deviation of 1.42. To compare the means of both groups, we ran a paired-samples t-test (Table 3). The t-statistic was 6.51, with df=11 (p < .01). The effect size for the difference between the groups was calculated with Cohen’s d (Table 4). The effect size was 1.88, which is a large effect size.

**Table 3.** Paired Samples Test for GDS and CBS-EOLC Scores Before and After Music Intervention in Residents and Caregivers.

| Outcome | Mean ± SD | Mean Difference (95% CI) | t | df | P value |
| --- | --- | --- | --- | --- | --- |
| **Geriatric Depression Rating Scale** |  |  |  |  |  |
| Before Music Therapy | 3.75 ± 1.75 | 3.00 (1.99 to 4.01) | 6.51 | 11.00 | <0.0001 |
| After Music Therapy | 0.75 ± 1.42 | | | | |
| **Caregiver Burden Scale** |  |  |  |  |  |
| Before Music Therapy | 23.00 ± 3.81 | 2.85 (1.29 to 4.40) | 3.98 | 12.00 | 0.0009 |
| After Music Therapy | 20.15 ± 3.05 | | | | |

**Table 4.**
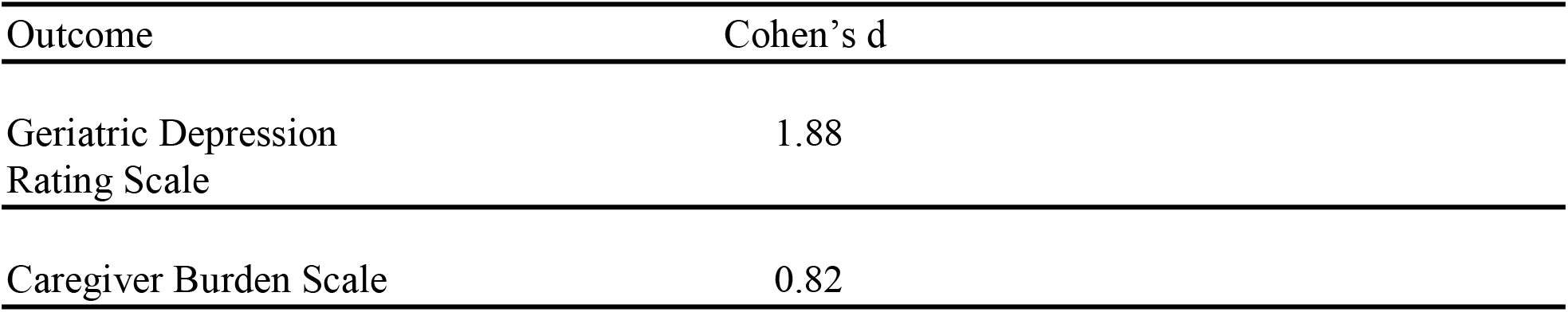
Calculated Effect Sizes for GDS and CBS-EOLC Scores Before and After Music Intervention.

The mean CBS-EOLC score for caregivers before the music intervention was 23, with a standard deviation of 3.81. The mean CBS-EOLC score for caregivers after the music intervention was 20.15, with a standard deviation of 3.05. A paired-samples t-test was run to compare the means of the two groups before and after intervention (Table 3). The t-statistic was 3.98, with df=12 (p < .01). The effect size was calculated with Cohen’s d (Table 4). The effect size was 0.82, which was a large effect size.

After the music intervention, caregivers also filled out a survey that included their experience as a caregiver, observations of the residents before and after the music performance, and perceptions of their own state of mind before and after the music. The average years of experience in working with patients in memory care was 11.5 years. All 13 caregivers reported that they noticed a shift in the resident’s mood and their own mood after the music performance. After the music performance, the post-music survey indicated that all patients that had previously been restless, became either content or indifferent, and the number of residents who were content increased. After the performance, caregivers who reported that they were “stressed”, “restless”, “exhausted”, and “tired”, had later reported being “comfortable”, “relaxed”, and “calm”.

## Discussion

This study has shown that receptive music therapy in a group setting can be used to help elderly people with Alzheimer’s Disease and other forms of dementia by decreasing depression symptoms. This approach can also be helpful to those who have moderate to mild cognitive impairment. Residents showed significant decreases in GDS scores after receiving the musical intervention compared to before. Effect sizes were large, indicating clinical relevance. Other studies point to corroborating results.^30–31,^ Caregiver burden scores were also significantly decreased after the music intervention. Effect sizes were also large. Another interesting finding was that despite a low proportion of residents who filled out the GDS, the rate of attendance was very high. Each performance retained this number of residents, which shows sustained engagement and appreciation for music. Music allows people to enter a tranquil and calm state of mind. The pleasant cumulation of different rhythms, pitches, and tones can elicit positive thoughts that may distract someone from more negative thoughts. During the performance, caregivers self-reported that many residents smiled while some cried. Music may also have the ability to remind the residents of different memories or allow them to express themselves, helping to bring balance to their emotions. Listening to music with a live audience and performance may also increase feelings of belongingness and social connectedness, which can be a protective factor to suicide and serve as a treatment for depressive and suicidal thoughts.^32^

There are some limitations to this study. This study lasted for a short period of time and did not assess the impact of music therapy in the long term. Limitations in resources prevented this study from being conducted at multiple residents and community centers, which may restrict the applicability and generalizability of this study to different settings. This also prevented data collection from a larger sample size. In future studies, it is recommended that researchers sample from a diverse background of patients in community, care centers, and mental health institutions to broaden understanding of music therapy impacts to those places. Future studies can also investigate whether different tempos, rhythms, and genres of music may contribute to varying effects on depression in dementia patients.

## Conclusion

In this study, we showed that after six 30-minute sessions of music therapy, there were significant improvements in depression in elderly residents with dementia and caregivers who worked with these patients at the facility. Music therapy may also be a cost-effective treatment for residents in this setting and for those with mild to moderate cognitive impairment. Music therapy should be integrated into residents’ and caregivers’ routines with regularly scheduled live performances to maximize treatment efficacy.

## Data Availability

All data produced in the present study are available upon reasonable request to the authors
and all data produced in the present work are contained in the manuscript

## Acknowledgments

We thank the staff for permitting and collaborating with this investigation and the residents at Amavida Living in Fort Myers Florida for their participation.

## CRediT authorship contribution statement

**Hieu Nguyen:** Writing – review & editing, Writing – original draft, Project administration, Methodology, Formal analysis, Data curation, Conceptualization. **Tomoteru Seki:** Writing – review & editing, Methodology, Formal analysis. **Wendy Chase:** Validation, Supervision.

